# Prepubertal Body Mass Index Reshapes the Distribution of Pubertal Growth Velocity in Vietnamese Children: A SITAR-Based Quantile Analysis

**DOI:** 10.64898/2026.09.04.26362313

**Authors:** Nhan T. Ho

## Abstract

**Background:** Prepubertal body mass index (BMI) has been reported to associate with pubertal growth spurts, but whether BMI reshapes the entire distribution of pubertal growth, not just its average, is unknown, particularly in Southeast Asian children.

**Methods:** We studied 8,018 boys, 5,495 girls from the 2018-2025 cohort of schoolchildren in 3 major cities in Vietnam with SITAR derived growth curves and prepubertal BMI z scores. Individual height SITAR size, timing, and intensity (velocity) parameters was modeled with linear quantile mixed models across quantiles 0.05 to 0.95, separately by sex, with a sensitivity model re-centered on each child’s own age at peak height velocity.

**Results:** In boys, the BMI to velocity association reversed across the distribution, from 0.045 cm/year per SD at the 5th percentile to −0.101 at the 95th (heterogeneity p<0.0001), and this reversal persisted after re-centering on each child’s own pubertal peak (p<0.0001). In girls, the association stayed positive at every quantile, from 0.132 to 0.187, without significant variation across the distribution in chronological age (heterogeneity p=0.22), though a significant distributional pattern reemerged once re-centered on pubertal peak (p=0.03). BMI compressed the spread of pubertal timing in both sexes and of intensity in girls (p<0.0001 for timing in both sexes, p=0.000002 for intensity in girls). Individual timing and intensity were inversely correlated in boys (r=-0.85) and positively correlated in girls (r=0.74).

**Conclusion:** Prepubertal BMI reshapes the distribution of pubertal growth in sex-specific ways. Clinical and epidemiological pediatrics should account for childhood BMI’s complex, distribution-specific, sex-dependent impact on adolescent development.

## INTRODUCTION

Childhood overweight and obesity have risen sharply across Vietnam over the past two decades, and our own analysis of urban Vietnamese children school health cohort recently documented an alarming pattern. More than 47% of boys and 26% of girls aged 5 to 18 years across three major Vietnamese cities now carry excess weight, several times higher than the World Health Organization threshold for a public health emergency ^1^. That same analysis showed something more specific and more puzzling. Children with obesity were taller than their peers in early childhood, but this advantage disappeared by the end of puberty, as if the pubertal growth spurt itself absorbed the early height gain ^1^. A companion analysis of this cohort further showed that body mass index reshapes the entire distribution of height for age, not just its mean, which raises the question of whether the pubertal growth spurt underlying that shift can be seen directly. What drives the convergence in final height, whether it reflects a shift in timing, a shift in intensity, or both, has not been examined in Vietnamese children.

International evidence on the timing question is extensive. A landmark study of nearly 157,000 Copenhagen children found that higher body mass index at age seven predicted an earlier onset of the pubertal growth spurt and earlier peak height velocity in both boys and girls ^2^. Multi-cohort United States data confirmed that faster weight gain in infancy and early childhood predicted a younger age at peak height velocity, with a stronger effect in boys ^3^, and a Boston birth cohort found that obesity as early as age two to four years was linked to earlier peak height velocity ^4^. Belarusian and American cohorts jointly showed that higher body mass index in middle childhood predicted both earlier puberty and slower subsequent linear growth ^5^. A large Swedish cohort added an important nuance, since the inverse relationship between body mass index and pubertal timing held only below a body mass index near the overweight threshold, and essentially disappeared above it in boys ^6^.

Beyond timing, prepubertal body mass index appears to reshape the pubertal growth spurt itself. In Chinese children, higher prepubertal body mass index predicted not only earlier peak height velocity but also a smaller peak height velocity in boys ^7^, and early peak height velocity combined with a small, short spurt independently predicted lower final height and higher late adolescent overweight risk ^8^. The effect appears dose dependent. Spanish children with moderate to extreme obesity showed progressively smaller pubertal height gains with increasing peak childhood body mass index ^9^, Japanese data from more than 13,000 children found that obesity blunted pubertal growth regardless of pubertal timing ^10^, and Swedish obesity treatment cohorts found the growth spurt diminished in class one obesity and essentially absent in class two obesity among boys ^11^. The biological pathways behind these patterns are only partly mapped. Genome wide studies have linked pubertal growth, pubertal timing, and childhood adiposity to shared genetic loci, but these loci do not fully explain the epidemiological associations, leaving the underlying mechanism only partly understood ^12^.

Evidence from Southeast Asia remains thin. The closest regional data come from Chinese cohorts already noted and from a Taiwanese study of schoolchildren, which found a more complicated pattern in which obese girls actually grew more slowly than underweight girls once puberty began, suggesting that puberty itself, not body mass index, dominates growth during that window ^13^.

There is also a methodological gap that cuts across every study reviewed above. Almost all of this literature, in every population and at every scale, compares mean age at peak height velocity or mean peak height velocity between body mass index groups, using linear regression or mixed effects models fit to the conditional average. That approach implicitly assumes body mass index shifts pubertal growth by roughly the same amount for every child, an assumption that is rarely tested and may not hold. If obesity disproportionately affects children who are already fast growers or already slow growers, a difference in group means could understate the true effect or miss it altogether. Quantile regression avoids this assumption by estimating the association between an exposure and an outcome separately at multiple points across the outcome’s distribution rather than only at its mean, and its extension to clustered and longitudinal data through quantile mixed models makes it possible to apply this idea to repeated growth measurements within the same child ^14,15^.

This paper addresses both gaps mentioned above. Using the annual school health check data from 2018 to 2025 of urban Vietnamese children, this paper applies quantile regression approach to SuperImposition by Translation And Rotation (SITAR) growth curve model derived measures of pubertal velocity, timing, and intensity, to test directly whether prepubertal body mass index widens, narrows, or shifts the entire distribution of pubertal growth, not only its average, in boys and girls, a question mean based methods in this field have not been able to answer.

## METHODS

This study performed retrospective analysis of de-identified data from annual school health check of students attending a private school system in three major cities in Vietnam (Hanoi, Ho Chi Minh City, and Haiphong), from 2018 to 2025.

Prepubertal body mass index was defined as in our earlier trajectory study ^16^, using the World Health Organization body mass index for age z score at each child’s earliest measurement taken at or before age 11 years for boys and age 9 years for girls, based on the WHO growth reference for school aged children and adolescents ^17^. Pubertal growth curves for the same children were already characterized using the sex-specific SITAR growth curve models in our companion paper ^16^ and the fitted SITAR models, the random effect estimates for every child, and the population age and magnitude of peak height velocity were carried forward for this paper without refitting. SITAR is a shape invariant mixed effects curve model that summarizes each child’s height trajectory through three random effects representing size, timing, and intensity of growth relative to a shared population curve ^18^. Baseline characteristics were compared across prepubertal body mass index categories separately within each sex, using the Kruskal-Wallis test for continuous variables and the chi-square test for categorical variables, since boys and girls differ substantially in both population pubertal timing and the sex composition of each body mass index category, and pooling them would confound any category comparison with sex.

Two outcome measures were derived from these existing fits. First, individual height velocity was estimated on a fixed six month age grid, from 8 to 16 years in boys and from 8 to 14.5 years in girls, by taking the central finite difference of each child’s model predicted height curve, using a one month step. The female grid was restricted to 14.5 years because the girls’ SITAR model lacks reliable data support beyond that age in this cohort, and extending it further extrapolated the fitted curve into a range with no real observations behind it. This produced a repeated measures panel linking velocity at each age point to prepubertal body mass index, sex, and hospital site. Second, each child’s three scalar SITAR random effects, capturing size, timing, and intensity, were treated as single cross sectional outcomes and modeled directly against prepubertal body mass index.

The central question was whether prepubertal body mass index shifts the entire distribution of pubertal velocity rather than only its mean. Linear quantile mixed models were fit to the velocity panel across quantiles from 0.05 to 0.95, with a random intercept for each child and fixed effects for prepubertal body mass index interacted with a natural cubic spline of age, adjusting for city ^14^. Models were fit with a gradient search algorithm for the asymmetric Laplace likelihood, refit at a higher iteration limit for any quantile that did not meet the algorithm’s convergence criterion, and, for any quantile that still failed to produce a fit, refit a final time with a derivative free algorithm as a fallback ^19^. Boys and girls were modeled separately given the different biology of pubertal timing between sexes. Confidence intervals came from a cluster level bootstrap that resampled children with replacement, and a joint Wald test compared the body mass index coefficient across quantiles to test formally for distributional heterogeneity ^20^. A parallel pooled model with a sex by body mass index interaction term checked whether stratifying by sex was statistically justified. As a sensitivity analysis, the same quantile model was refit with velocity expressed relative to each child’s own age at peak height velocity instead of chronological age, to separate effects on growth magnitude from effects on growth timing.

The three scalar SITAR parameters were analyzed with quantile regression, again by sex, using the same quantile grid and bootstrap approach ^15^. An exploratory mediation analysis tested whether pubertal timing mediated the path from prepubertal body mass index to late adolescent height for age z score, using a product of coefficients approach with a percentile bootstrap confidence interval for the indirect effect ^20,21^. Additional sensitivity checks varied the spline degrees of freedom, excluded one hospital site at a time, and compared children with and without sufficient visits for SITAR fitting.

All analyses were performed in R version 4.5.1 ^22^ using the sitar ^18^, lqmm ^23^, quantreg ^24^, and nnet ^25^ packages, with parallel computation for the bootstrap procedures using the furrr ^26^ and future ^27^ packages. Statistical significance was set at a two sided p value below 0.05.

## RESULTS

### Study sample

Of the 20,082 children (10,848 boys, 9,234 girls) with sufficient visits for a SITAR growth curve, 13,513 (8,018 boys, 5,495 girls) also had a valid prepubertal BMI z score and were included in the distributional analyses. The population age at peak height velocity (APHV) was 12.2 years (95% CI 12.20 to 12.21) in boys and 9.2 years (95% CI 9.18 to 9.22) in girls, with corresponding peak height velocity (PHV) of 9.34 cm per year (95% CI 9.33 to 9.36) and 7.00 cm per year (95% CI 6.98 to 7.02) (**Figure 1**).

**Figure 1.**
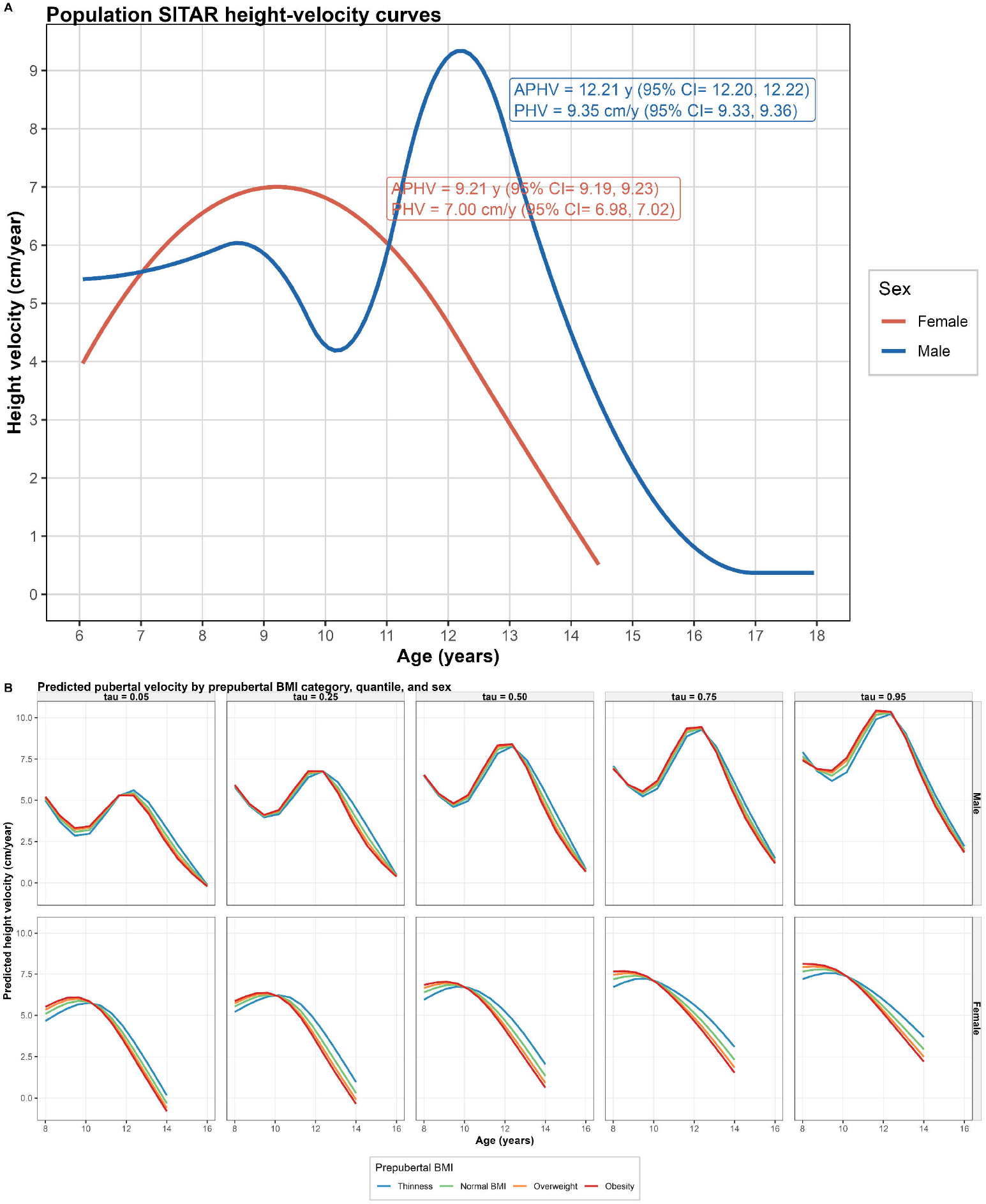
Population SITAR height velocity curves, by sex and Predicted pubertal velocity by prepubertal BMI category and quantile, by sex. A. Population average height velocity curve for boys and girls, reused from the companion SITAR growth curve analysis without refitting. Age in years on the x axis, height velocity in centimeters per year on the y axis. Blue is boys, red is girls, each annotated with its own age at peak height velocity (APHV) and peak height velocity (PHV) with 95 percent confidence intervals. Boys show a minor early peak around age 8 to 9 years followed by the adolescent growth spurt peaking at 12.2 years. Girls show a single, earlier, comparatively lower peak at 9.2 years. The female curve is shown only through age 14.5 years, since the fitted SITAR model does not have reliable data support beyond that age in girls in this cohort, and extending the curve further would extrapolate past the data. This figure establishes the population backdrop against which the individual level, BMI stratified curves should be read. B. Each panel shows predicted height velocity across age for one sex, in rows, and one quantile from 0.05 to 0.95, in columns. Within each panel, lines show prepubertal BMI category, thinness, normal BMI, overweight, and obesity, from the sex-stratified quantile mixed models. Boys are shown from age 8 to 16 years, girls from age 8 to 14.5 years, reflecting the reliable data range of each sex-specific SITAR model. In boys, the ordering of BMI categories reverses between the lowest and highest quantile panels, with the obesity curve above the others at the 0.05 quantile and below the others at the 0.95 quantile. In girls, the obesity curve stays at or above the other BMI categories across all five quantile panels.

Prepubertal BMI category was unevenly distributed by sex. Boys made up 59.3 percent of the sample overall but 83.0 percent of the obesity group, while girls made up 40.7 percent overall but only 17.0 percent of the obesity group (**Table 1**).

**Table 1.** Baseline characteristics of the analytic cohort by sex and prepubertal BMI category.

|  | Male |  |  |  |  |  | Female |  |  |  |  |  |
| --- | --- | --- | --- | --- | --- | --- | --- | --- | --- | --- | --- | --- |
|  | Thinness<br>(N=193) | Normal<br>BMI<br>(N=3613) | Overweight<br>(N=1862) | Obesity<br>(N=2350) | Total<br>(N=8018) | p value | Thinness<br>(N=180) | Normal<br>BMI<br>(N=3648) | Overweight<br>(N=1186) | Obesity<br>(N=481) | Total<br>(N=5495) | p value |
| Age at baseline, years, median (Q1 to Q3) | 7.300 (6.300, 9.100) | 7.000 (6.400, 8.600) | 8.400 (6.800, 10.100) | 8.200 (6.900, 9.700) | 7.900 (6.500, 9.500) | < 0.001 | 7.100 (6.300, 8.100) | 7.000 (6.400, 8.025) | 7.400 (6.400, 8.300) | 7.200 (6.300, 8.200) | 7.100 (6.300, 8.200) | < 0.001 |
| Individual APHV, years, median (Q1 to Q3) | 12.311 (12.037, 12.566) | 12.463 (12.140, 12.712) | 12.184 (11.832, 12.489) | 12.096 (11.724, 12.422) | 12.229 (11.897, 12.519) | < 0.001 | 9.199 (8.581, 9.870) | 9.116 (8.460, 9.900) | 9.193 (8.682, 9.787) | 9.256 (8.671, 9.816) | 9.199 (8.607, 9.852) | 0.481 |
| SITAR timing (b), median (Q1 to Q3) | 0.101 (-0.173, 0.356) | 0.253 (-0.070, 0.502) | -0.026 (-0.378, 0.279) | -0.114 (-0.486, 0.212) | 0.019 (-0.313, 0.309) | < 0.001 | -0.011 (-0.629, 0.660) | -0.094 (-0.750, 0.690) | -0.017 (-0.528, 0.577) | 0.046 (-0.539, 0.606) | -0.011 (-0.603, 0.642) | 0.481 |
| SITAR intensity (c), median (Q1 to Q3) | -0.012 (-0.055, 0.033) | -0.032 (-0.078, 0.019) | 0.007 (-0.041, 0.054) | 0.017 (-0.030, 0.067) | 0.000 (-0.046, 0.049) | < 0.001 | -0.004 (-0.071, 0.063) | -0.042 (-0.090, 0.030) | 0.016 (-0.039, 0.072) | 0.032 (-0.025, 0.088) | 0.005 (-0.062, 0.068) | < 0.001 |
| Individual PHV, cm per year, median (Q1 to Q3) | 9.229 (8.850, 9.661) | 9.054 (8.642, 9.526) | 9.412 (8.973, 9.862) | 9.504 (9.073, 9.993) | 9.349 (8.925, 9.813) | < 0.001 | 6.972 (6.520, 7.459) | 6.713 (6.398, 7.211) | 7.111 (6.733, 7.526) | 7.229 (6.826, 7.644) | 7.033 (6.579, 7.494) | < 0.001 |
| Study site |  |  |  |  |  | < 0.001 |  |  |  |  |  | 0.001 |
| Hanoi, n (%) | 2398 (66.4%) | 102 (52.8%) | 1261 (67.7%) | 1507 (64.1%) | 5268 (65.7%) |  | 2445 (67.0%) | 105 (58.3%) | 755 (63.7%) | 295 (61.3%) | 3600 (65.5%) |  |
| Hochiminh city, n (%) | 885 (24.5%) | 76 (39.4%) | 454 (24.4%) | 606 (25.8%) | 2021 (25.2%) |  | 894 (24.5%) | 65 (36.1%) | 333 (28.1%) | 136 (28.3%) | 1428 (26.0%) |  |
| Haiphong, n (%) | 330 (9.1%) | 15 (7.8%) | 147 (7.9%) | 237 (10.1%) | 729 (9.1%) |  | 309 (8.5%) | 10 (5.6%) | 98 (8.3%) | 50 (10.4%) | 467 (8.5%) |  |

*BMI (body mass index) category is based on the World Health Organization body mass index for age z score at the earliest measurement occurring at or before age 11 years for boys or age 9 years for girls. APHV, age at peak height velocity. PHV, peak height velocity. Baseline characteristics were compared across prepubertal body mass index categories separately within each sex, using the Kruskal-Wallis test for continuous variables and the chi-square test for categorical variables.*

### Prepubertal BMI reshapes the velocity distribution differently by sex

In the primary quantile model, prepubertal BMI z score showed a clear, sex-specific pattern across the velocity distribution. In boys, the association with velocity started positive at the low end of the distribution (0.045 cm per year per SD at the 5th percentile) and crossed to negative at the high end (−0.101 cm per year per SD at the 95th percentile), a highly significant departure from a single shared effect (heterogeneity Wald p<0.0001). In girls, the association was positive across every quantile examined, ranging from 0.132 to 0.187 cm per year per SD, but unlike boys this did not vary significantly across the distribution (heterogeneity Wald p=0.215). A formal test of a sex by BMI interaction across the pooled model was strongly significant (p<0.0001), confirming that boys and girls respond differently even though girls’ own distributional heterogeneity did not reach significance (**Figure 1, Figure 2, Table 2, Table S1, Table S2**).

**Table 2.**
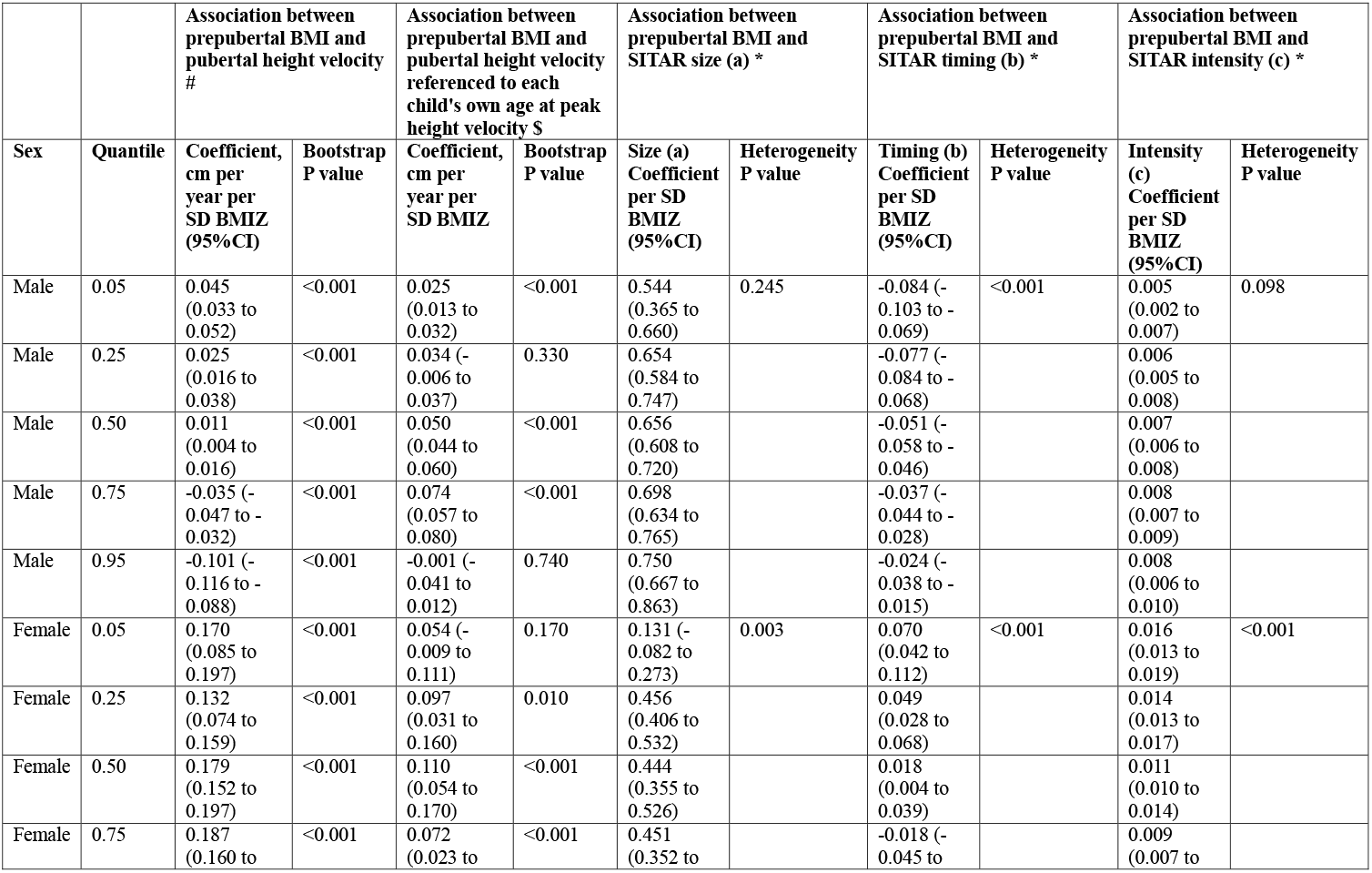
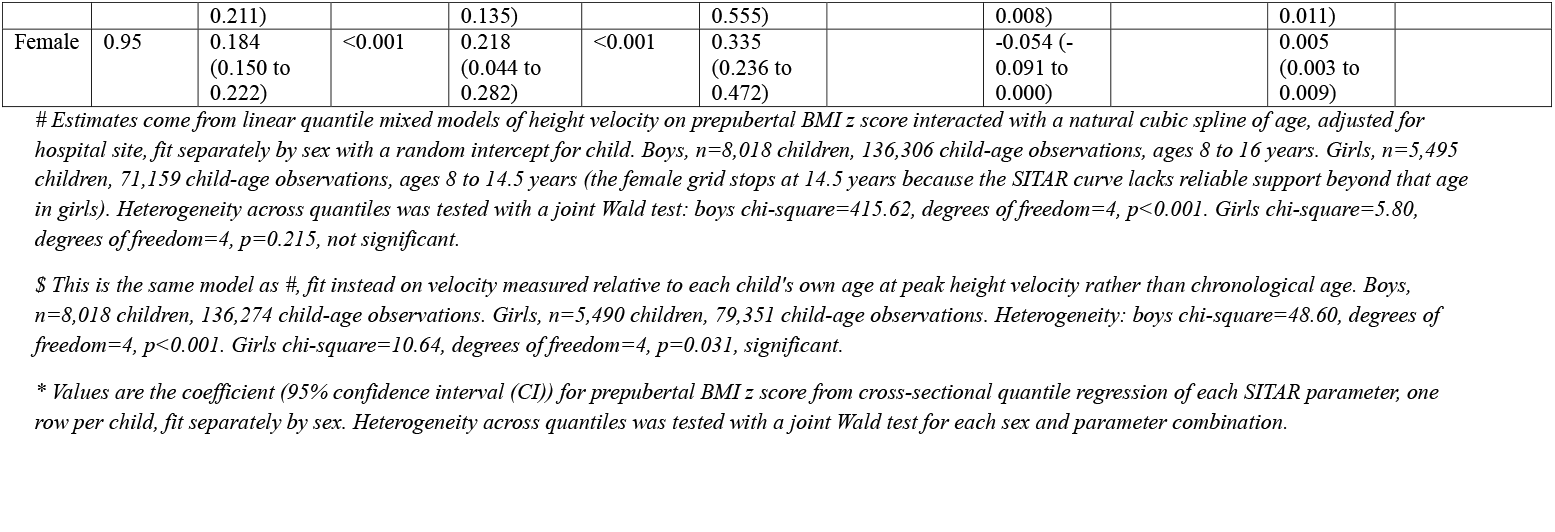
Association between prepubertal BMI and pubertal height velocity and SITAR parameters across quantiles, by sex.

**Figure 2.**
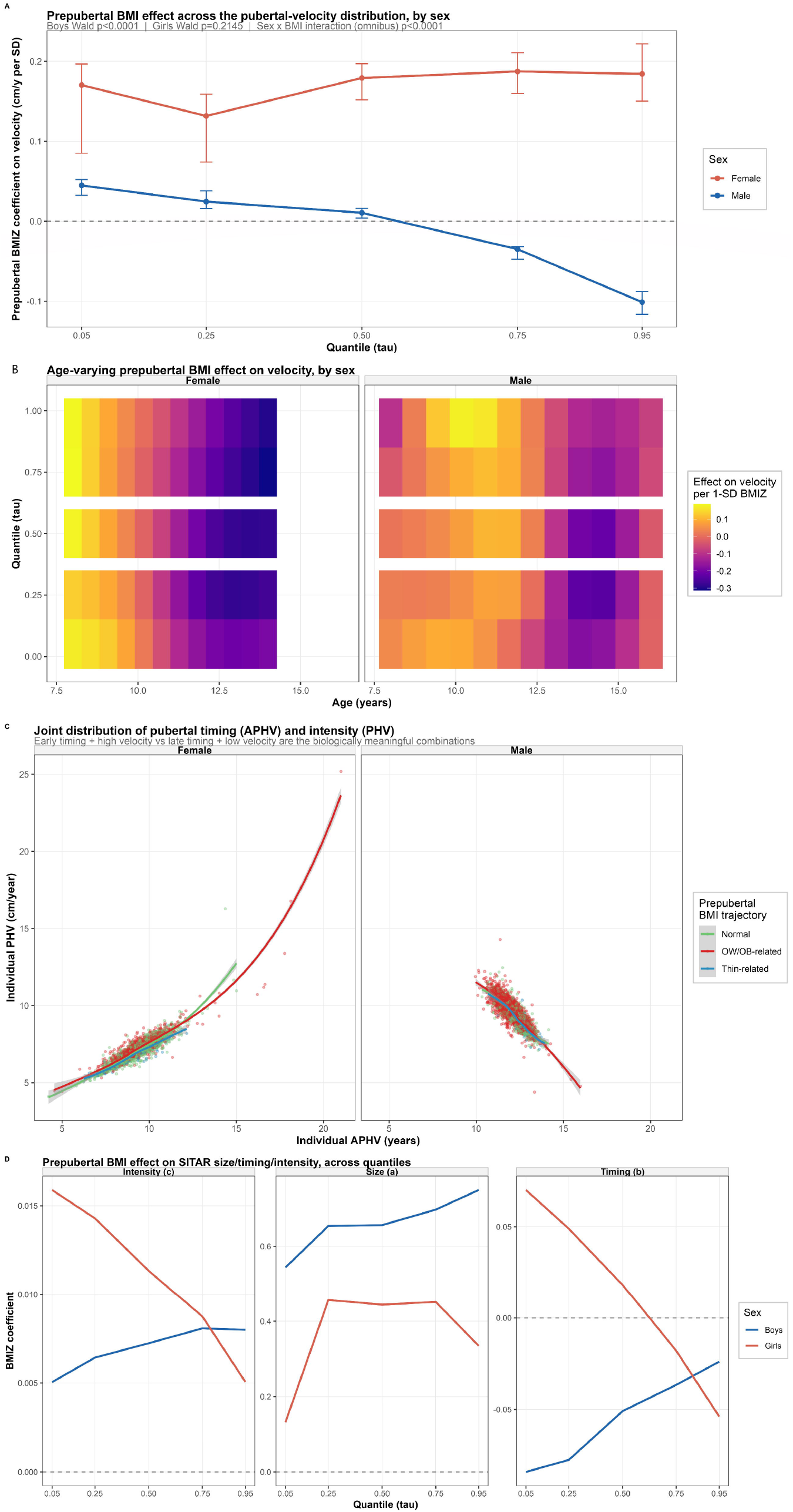
Prepubertal BMI effect across the pubertal velocity distribution, joint distribution of pubertal timing and intensity, and prepubertal BMI effect on SITAR size, timing, and intensity parameters across quantiles, by sex. A. Coefficient for prepubertal BMI z score on velocity, in centimeters per year per standard deviation, against quantile from 0.05 to 0.95, boys in blue, girls in red, with 95 percent bootstrap confidence intervals. The dashed line marks no effect. The subtitle reports the heterogeneity Wald test for each sex (boys p<0.0001, girls p=0.215, not significant) and the joint sex by BMI interaction test (p<0.0001). B. Heat maps of the estimated effect of a one standard deviation increase in prepubertal BMI z score on velocity, by age and quantile, for each sex. Warmer colors indicate a positive effect, cooler colors a negative effect. Both heat maps are now fully estimated across the full quantile range in both sexes. C. Each point represents one child, plotted by individual age at peak height velocity on the x axis and individual peak height velocity on the y axis, with girls on the left and boys on the right. Points are colored by prepubertal BMI trajectory, defined from the earliest and latest prepubertal BMI category recorded for that child, as normal, thinness related, or overweight or obesity related. Curved lines show a loess smooth with a shaded 95 percent confidence band for each trajectory group. As the subtitle notes, the biologically meaningful contrast is between early timing paired with high velocity and late timing paired with low velocity. The opposite slope of the smoothed lines between the girls’ panel and the boys’ panel reflects the opposite sign correlation. D. Three panels show the coefficient for prepubertal BMI z score, from the cross-sectional quantile regressions, plotted against quantile from 0.05 to 0.95, for intensity on the left, size in the middle, and timing on the right. Boys are shown in blue and girls in red. The dashed horizontal line marks no effect. For timing, the girls’ line crosses from positive to negative across the quantile range, while the boys’ line stays negative but shrinks in magnitude, both patterns consistent with prepubertal BMI narrowing the spread of pubertal timing rather than shifting it uniformly. For intensity, girls show a similar narrowing pattern, while boys show a smaller and only borderline significant version of the same pattern.

### Sensitivity analysis anchored to each child’s own pubertal peak

When velocity was re-expressed relative to each child’s own age at peak height velocity rather than chronological age, the effect of BMI still varied significantly across quantiles in both sexes. In boys, associations were significant and positive at the 5th percentile and from the median through the 75th percentile (0.025 to 0.074 cm per year per SD), but not at the 25th or 95th percentiles (heterogeneity Wald p<0.0001). In girls, associations were significant at every quantile except the 5th percentile, ranging from 0.072 to 0.218 cm per year per SD, and heterogeneity across quantiles was also significant (heterogeneity Wald p=0.031). This indicates that BMI reshapes the velocity distribution in both sexes even after accounting for each child’s own pubertal timing, so the distributional pattern seen in chronological age is not simply an artifact of when the growth spurt occurs (**Table 2, Figure 3**).

**Figure 3.**
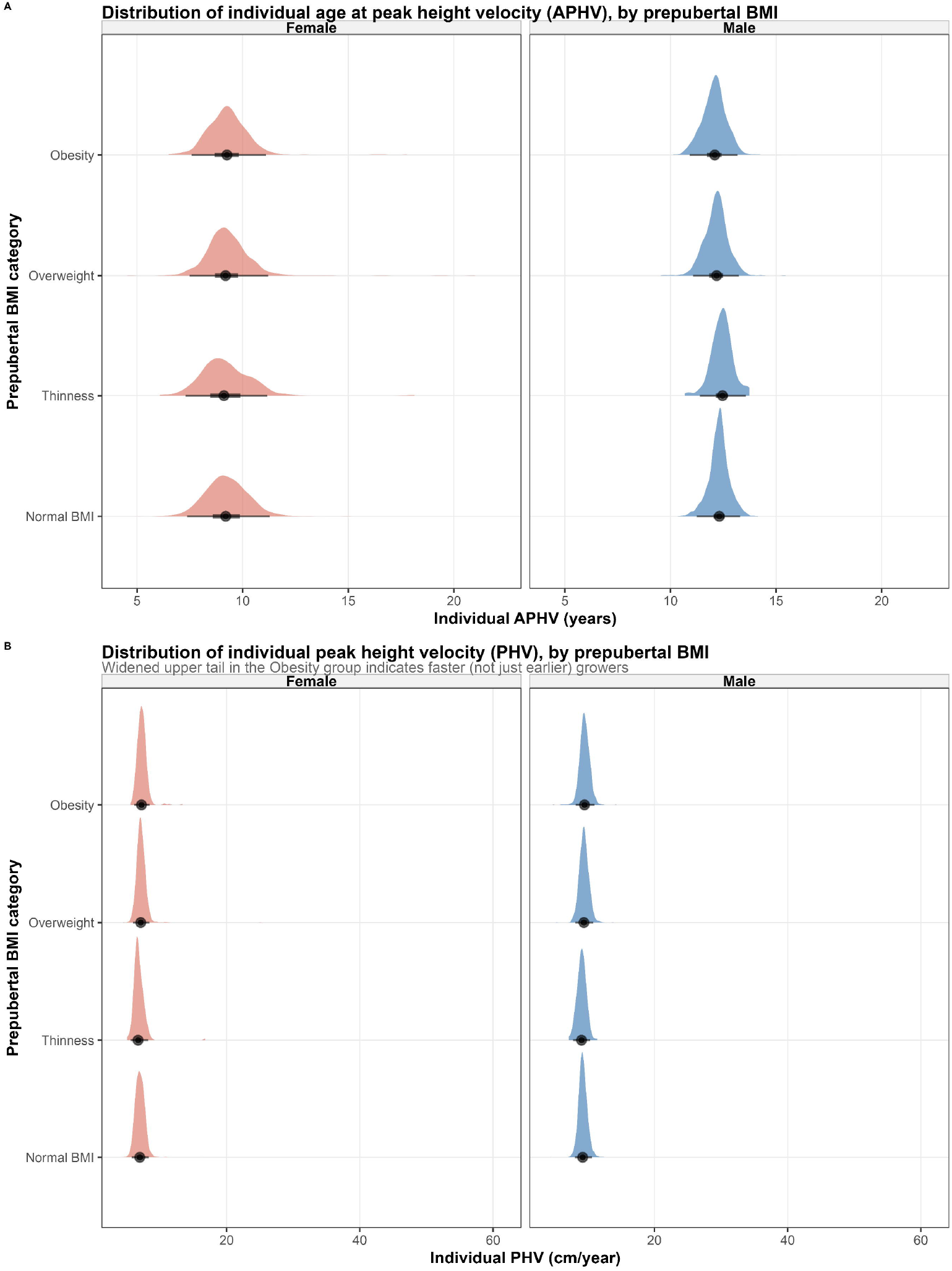
Distribution of individual age at peak height velocity and peak height velocity, by prepubertal BMI category and sex. A. Each half violin and point interval shows the distribution of individual age at peak height velocity, in years, for one prepubertal BMI category, separately for girls on the left and boys on the right. The dot marks the median and the horizontal line marks the interquartile range. Distributions are visibly narrower and shifted earlier in boys than in girls, consistent with the difference in population timing. B. Each half violin and point interval shows the distribution of individual peak height velocity, in centimeters per year, for one prepubertal BMI category, separately for girls on the left and boys on the right. As the subtitle notes, the obesity category shows a visibly wider upper tail than the other BMI categories in both sexes, indicating that some children with obesity have a substantially faster growth spurt than the typical child, not simply an earlier one.

### BMI reshapes SITAR size, timing, and intensity parameters differently

Quantile regression on the three scalar SITAR parameters showed a broadly consistent pattern across both sexes for pubertal timing and intensity, best described as a narrowing of the tails. For timing, boys showed a negative BMI coefficient at every quantile, but its magnitude shrank from −0.084 at the 5th percentile to −0.024 at the 95th (Wald p<0.0001), meaning BMI advanced timing most among boys who were already the earliest developers. Girls showed a coefficient that flipped sign, from +0.070 at the 5th percentile to −0.054 at the 95th (Wald p<0.0001), meaning higher BMI pulled the earliest developers later and the latest developers earlier, compressing the spread of pubertal timing overall. A similar compression appeared for intensity in girls, where the BMI coefficient fell from 0.016 at the 5th percentile to 0.005 at the 95th (Wald p=0.000002), and a smaller, borderline effect in the opposite direction appeared in boys (0.005 to 0.008, Wald p=0.098). For the size parameter, BMI’s association was positive across all quantiles in both sexes and did not vary significantly by quantile in boys (Wald p=0.245), while girls showed significant but less orderly heterogeneity (Wald p=0.003) (**Figure 2, Table 2**).

### Timing and intensity move in opposite directions by sex

The correlation between individual APHV and individual PHV was strongly negative in boys (r=-0.85, n=8,018) and strongly positive in girls (r=0.74, n=5,495). In practical terms, boys who entered puberty earlier tended to have a more intense growth spurt, while girls who entered puberty earlier tended to have a less intense one. This opposite-signed relationship held within every prepubertal BMI trajectory group (**Figure 2, Table S3**).

### Exploratory mediation through pubertal timing

Using each child’s last observed height for age z score at or after age 15 as a proxy for late-adolescent height, prepubertal BMI was associated with earlier pubertal timing in both sexes (path a: −0.094 in boys, −0.058 in girls), and earlier timing was in turn associated with a lower proxy late-adolescent height for age z score (path b: 0.072 in boys, 0.499 in girls). The resulting indirect effects were small and not statistically significant, at −0.0067 (95 percent CI −0.0126 to 0.0002) in boys and −0.0291 (95 percent CI −0.0863 to 0.0141) in girls, the latter based on only 152 girls with both exposure and outcome data available. These results should be treated as exploratory given the proxy outcome and limited sample size, particularly in girls (**Table S4**).

Of note, sensitivity analysis of the median velocity model to spline degrees of freedom showed the girls’ estimate was stable across specifications. The boys’ estimate at the median was more sensitive to spline flexibility, which should be kept in mind when interpreting the boys’ median result specifically (**Table S5**). Leave-one-city-out sensitivity analysis of the median velocity model showed that estimates were consistent regardless of which hospital site was excluded, suggesting the primary findings are not driven by any single site (**Table S6**). Children who had enough visits to support a SITAR growth curve had a somewhat higher mean BMI z score than those who did not, in both sexes, which should be considered when generalizing these results to the full cohort (**Table S7**).

## DISCUSSION

This study found that prepubertal BMI does not simply shift the mean of pubertal growth velocity in Vietnamese children, it reshapes the distribution itself, and it does so differently in boys and girls. In boys, higher BMI predicted faster velocity among the slowest growers but slower velocity among the fastest growers, a pattern that persisted even after re-centering age on each child’s own peak. In girls, higher BMI was associated with faster velocity across nearly the entire distribution, and this association remained significant even after accounting for each girl’s own pubertal timing, though the shape of that association across quantiles was itself more uniform in girls than in boys. These findings extend our earlier observation in this same cohort that children with obesity lose their early height advantage by the end of puberty ^1^, by showing that this convergence is not driven by a single average slowdown but by a genuine narrowing, or in boys a partial reversal, of who grows fastest and who grows slowest.

The literature on BMI and pubertal timing is large and largely mean-based. Landmark cohorts from Denmark, the United States, and China have consistently linked higher prepubertal BMI to earlier peak height velocity ^2–4,28^, and cohorts following children into obesity treatment or severe obesity have shown blunted growth spurts with increasing BMI severity ^7–9,11^. Our SITAR-derived timing and intensity parameters agree with the general direction of this literature but add a layer these mean-based methods cannot capture. Rather than a uniform advance in timing, we found that BMI compressed the spread of pubertal timing in both sexes, pulling the earliest developers later and the latest developers earlier, most clearly in girls. A Swedish cohort previously reported that the BMI to timing association held only below the overweight threshold and vanished above it in boys ^6^, a nonlinearity that is consistent with a compression pattern rather than a straight-line shift, and our quantile results make that kind of nonlinearity visible directly rather than inferring it from subgroup cutoffs.

The clearest sex difference in our results was the opposite-signed correlation between individual timing and individual intensity, negative in boys and positive in girls. This has not, to our knowledge, been reported before. Genome-wide studies have linked pubertal timing, pubertal growth, and childhood adiposity to overlapping but not identical genetic loci ^12^, and a comprehensive recent review has emphasized that the hormonal pathways linking adiposity to puberty, particularly leptin signaling, act differently in boys and girls ^29^. A plausible interpretation is that in girls, adiposity works partly through the same pathway that governs timing itself, so earlier timing and higher intensity travel together, while in boys the growth-promoting and timing-advancing effects of adiposity may act through more separable pathways. This is a hypothesis our data can describe but not test directly.

Our exploratory mediation analysis, linking prepubertal BMI to a proxy of late-adolescent height through pubertal timing, found small indirect effects that did not reach statistical significance, and the girls’ estimate rested on only 152 children with both exposure and outcome data. This should be read as a preliminary signal rather than evidence of a specific pathway.

A regional study from Taiwan found that among girls, pubertal status appeared to matter more than BMI itself in shaping growth velocity ^13^. Our results only partly align with that observation. We did find that girls’ velocity distribution was more uniformly shifted by BMI than boys’, consistent with a less complex, more magnitude-driven effect in girls. However, this shift was not simply a byproduct of pubertal timing, since it remained statistically significant even after re-centering velocity on each girl’s own age at peak height velocity. This suggests that in our cohort, BMI has a genuine effect on growth magnitude in girls that coexists with, rather than substitutes for, its effect on timing, a distinction the Taiwanese study’s design could not directly address.

This study has real limitations. It is observational, drawn from a single Vietnamese cohort with substantially more boys than girls in the obesity category, and its mediation outcome is a proxy rather than true adult height. Strengths include a large sample, individual-level SITAR curves rather than group means, and a quantile modeling approach built specifically to detect distributional change. These results motivate our planned companion analysis in a separate paper regarding pubertal growth phenotypes in this same study cohort.

In brief, prepubertal BMI reshapes the distribution of pubertal growth in sex-specific ways that a single average curve cannot capture. Clinical and public health pediatrics must take into account childhood BMI’s complex, distribution-specific, sex-dependent impact on adolescent development.

## DECLARATION

### Data Sharing Statement

R codes are available from the corresponding author upon reasonable request. Individual patient-level data cannot be shared due to applicable privacy regulations and the terms of the institutional ethics approval.

### Funding statement

This study did not receive funding.

### Conflict of intertest

The author states that there is no conflict of interest.

### Use of Artificial Intelligence

The author performed all original research work regarding scientific content, analyses, interpretations and manuscript writing. The author used AI-assisted tools for language editing and grammar checking during manuscript preparation.

## Ethic and Consent statement

This study was approved by Vinmec Ethical Committee (approval number 0231/2024/CN/HDDD VMEC) with a waiver of individual informed consent as the study used de-identified routinely collected retrospective health data from school health examinations.

## Contributor’s statement

Nhan Thi Ho did conceptualization, data curation, formal analysis, investigation, methodology, project administration, resources, software, supervision, validation, visualization, writing original draft, and writing review & editing.

## Implications and Contribution

This study is the first to apply distributional, quantile-based methods to pubertal growth in Southeast Asian children, showing that prepubertal obesity reshapes growth velocity, timing, and intensity distributions differently by sex rather than simply shifting averages, filling a regional evidence gap and identifying sex-specific pathways for future mechanistic and clinical investigation.

## Notes

### Competing Interest Statement

The authors have declared no competing interest.

