## Supplementary Tables for "Prepubertal Body Mass Index Reshapes the Distribution of Pubertal Growth Velocity in Vietnamese Children: A SITAR-Based Quantile Analysis"

**Table S1. Formal test of a sex by prepubertal BMI interaction in the quantile model of pubertal velocity**

| **Quantile** | **Interaction coefficient (female minus male)** | **95% CI** |
| --- | --- | --- |
| 0.05 | -0.071 | -0.118 to -0.028 |
| 0.25 | -0.043 | -0.102 to -0.019 |
| 0.50 | -0.094 | -0.103 to -0.085 |
| 0.75 | -0.096 | -0.108 to -0.084 |
| 0.95 | -0.094 | -0.134 to -0.021 |

*From a pooled model of both sexes with a scalar sex by BMI interaction term and separate age spline by sex, fit at each headline quantile. This scalar interaction term represents the age-averaged difference in slope and is not directly comparable to the age-varying, sex-stratified curves in Figure 3. The joint Wald test across all five quantiles was strongly significant (p<0.001), supporting the decision to fit boys and girls in fully separate models.*

**Table S2. Convergence and fitting method summary for the linear quantile mixed models**

| **Pipeline** | **Quantile** | **Converged** | **Method used** |
| --- | --- | --- | --- |
| Boys velocity, primary | 0.05 | No | Gradient search |
| Boys velocity, primary | 0.25 | Yes | Gradient search |
| Boys velocity, primary | 0.50 | Yes | Gradient search |
| Boys velocity, primary | 0.75 | Yes | Gradient search |
| Boys velocity, primary | 0.95 | No | Gradient search |
| Girls velocity, primary | 0.05 | No | Gradient search |
| Girls velocity, primary | 0.25 | Yes | Gradient search |
| Girls velocity, primary | 0.50 | Yes | Gradient search |
| Girls velocity, primary | 0.75 | No | Gradient search |
| Girls velocity, primary | 0.95 | Yes | Derivative free |

*A model marked "No" under Converged still produced a usable coefficient in most cases, it simply did not satisfy the strict internal convergence counter, and the resulting estimate is retained. Rows for the age since APHV sensitivity models are omitted here for space but are available on request, none of them affected the reported estimates.*

**Table S3. Correlation between individual age at peak height velocity and individual peak height velocity, by sex**

| **Sex** | **Correlation (r)** | **N** |
| --- | --- | --- |
| Female | 0.742 | 5,495 |
| Male | -0.846 | 8,018 |

*Girls who entered puberty earlier tended to have a lower peak height velocity, while boys who entered puberty earlier tended to have a higher one.*

**Table S4. Exploratory mediation analysis of prepubertal BMI, pubertal timing, and late-adolescent height**

| **Sex** | **N** | **Path a, BMI to timing (b)** | **Path b, timing (b) to late HAZ** | **Indirect effect** | **95% CI** | **Direct effect (c prime)** |
| --- | --- | --- | --- | --- | --- | --- |
| Male | 1,249 | -0.094 | 0.072 | -0.007 | -0.013 to 0.000 | 0.029 |
| Female | 152 | -0.058 | 0.499 | -0.029 | -0.086 to 0.014 | 0.039 |

*HAZ, height for age z score, using each child's last observed value at or after age 15 years as a proxy for late-adolescent height. Indirect effect is the product of path a and path b, with a percentile bootstrap confidence interval. This analysis is exploratory. It uses a proxy outcome rather than measured adult height, and the female sample is small.*

**Table S5. Sensitivity of the median velocity model to spline degrees of freedom**

| **Sex** | **Spline degrees of freedom** | **Coefficient at the median** |
| --- | --- | --- |
| Boys | 3 | -0.006 |
| Boys | 4 | 0.011 |
| Boys | 5 | 0.065 |
| Girls | 3 | 0.173 |
| Girls | 4 | 0.177 |
| Girls | 5 | 0.172 |

*The primary model used 4 degrees of freedom for the age spline. The girls' estimate was stable across specifications. The boys' estimate at the median was more sensitive to spline flexibility, which should be kept in mind when interpreting the boys' median result specifically.*

**Table S6. Leave-one-city-out sensitivity analysis of the median velocity model**

| **Sex** | **City excluded** | **Coefficient at the median** |
| --- | --- | --- |
| Male | HHN | 0.011 |
| Female | HHN | 0.165 |
| Male | HCP | 0.009 |
| Female | HCP | 0.186 |
| Male | HHP | 0.011 |
| Female | HHP | 0.177 |

*Estimates were consistent regardless of which hospital site was excluded, suggesting the primary findings are not driven by any single site.*

**Table S7. Comparison of children with and without sufficient visits for SITAR fitting**

| **Sex** | **SITAR eligible** | **N** | **Mean BMIZ, all visits** |
| --- | --- | --- | --- |
| Male | No | 40,099 | 1.13 |
| Male | Yes | 10,848 | 1.40 |
| Female | No | 36,754 | 0.35 |
| Female | Yes | 9,234 | 0.47 |

*Children who had enough visits to support a SITAR growth curve had a somewhat higher mean BMI z score than those who did not, in both sexes, which should be considered when generalizing these results to the full cohort.*
